# Detection of Spreading Depolarization with Noninvasive Electroencephalography

**DOI:** 10.64898/2026.10.28.26365092

**Authors:** Alireza Chamanzar, Morteza Zabihi, Sebastian Major, Shravan Sivakumar, Jonathan Elmer, Lori Shutter, Aman B. Patel, Christopher J. Stapleton, Jens P. Dreier, Pulkit Grover, Edilberto Amorim, David Y. Chung, Eric S. Rosenthal

## Abstract

**Background and Purpose:** Spreading depolarizations (SDs) are waves of near-complete breakdown of the transmembrane ion gradients that underlie migraine aura and occur in patients with moderate-to-severe traumatic, ischemic or hemorrhagic brain injury. SDs are involved in primary and secondary brain injury. They are associated with poor neurological outcome and represent a modifiable therapeutic target. However, despite decades of investigation, SD detection still requires invasive subdural electrocorticography (ECoG) using an implanted subdural electrode strip, restricting monitoring to a small subset of neurosurgical patients.

**Methods:** We developed WAVEFRONT, a noninvasive scalp electroencephalography (EEG) biomarker of SD based on spatiotemporal propagation characteristics of delta-band EEG activity. The biomarker was trained and validated using concurrent, multi-day scalp EEG and invasive subdural electrocorticography recordings from patients with traumatic brain injury (TBI), subarachnoid hemorrhage (SAH), or malignant hemispheric stroke (MHS) across three institutions. We assessed out-of-sample performance across institutions, diagnosis, and surgical approaches, as well as calibration, temporal synchrony, and spatial specificity relative to gold-standard SDs annotated on ECoG. Clinical utility was further evaluated in 607 patients comatose after cardiac arrest from the multicenter International Cardiac Arrest REsearch (I-CARE) consortium, a population in whom invasive monitoring is not performed. Outcome analyses excluded patients who died (Cerebral Performance Category [CPC] 5) because death may reflect decisions to withdraw life-sustaining treatment in addition to neurological injury.

**Results:** A total of 1,119 gold-standard SDs were annotated in 32 patients (TBI, n=17; SAH, n=12; MHS, n=3; range 0–187 SDs per patient). Across exhaustive cross-validation and institution-, diagnosis-, and surgical approach-based hold-out analyses, WAVEFRONT models trained on one set of patients consistently maintained high performance when evaluated in previously unseen out-of-group cohorts. At the optimal operating point, WAVEFRONT achieved a mean cross-validated true positive rate of 97.3±1% with a false positive rate of 6.0±0.5%, calculated over 2-min sliding windows with 30-s overlaps. The cumulative duration of WAVEFRONT detections within 30-hour windows strongly correlated with the number of SDs measured by invasive ECoG (R^2^=0.7411). Scalp-detected depressions were spatially consistent with the location of the invasive recording strip, and visual inspection demonstrated close temporal correspondence with invasive detections, including during SD clusters. External validation in the independent I-CARE cohort using WAVEFRONT trained on the invasive-monitoring cohort demonstrated significantly greater SD burden on day 3 of post-arrest EEG monitoring among patients with poor neurological outcome (CPC 3-4, n=29) than those with good outcome (CPC 1-2, n=225; p=0.007).

**Conclusions:** Eight decades after Leão’s description of SD in 1944, we propose and validate a noninvasive scalp EEG biomarker of SD. Consistent performance across institution-, diagnosis-, and surgical approach-based hold-out analyses, together with burden calibration, spatial specificity, and temporal synchrony with ECoG detections, supports the validity of the WAVEFRONT framework. The association of biomarker burden with neurologic outcome after cardiac arrest suggests that noninvasive monitoring may enable investigation of spreading depolarizations in populations where invasive monitoring is not feasible. Future studies should evaluate prospective performance, treatment responsiveness, and disease-specific associations of the SD biomarker with clinical outcome.

## Introduction

Here, we validate a noninvasive scalp electroencephalography (EEG) biomarker of spreading depolarizations (SDs) against simultaneous intracranial recordings in a multicenter cohort of craniotomy and craniectomy patients. First defined in 1944 by Leão [1], SDs are pathological waves of abrupt, near-complete breakdown of transmembrane ion gradients that propagate slowly (1-8 mm/min) across the cortex and disrupt cortical activity [2–5]. SDs can cause acute injury in the presence of metabolic stress in the tissue through which they propagate [6–8]. Over the past two decades, invasive human recordings have established SDs as fundamental mechanisms of primary and secondary injury in major acute brain disorders, including traumatic brain injury (TBI), subarachnoid hemorrhage (SAH), and ischemic and hemorrhagic stroke [3,9–14]. SDs are strongly associated with progressive lesion expansion, delayed cerebral ischemia, and poor neurological outcomes [9,15–19]. Preclinical and clinical data suggest that anesthetics (e.g., ketamine [20–26]), vasodilators (e.g., cilostazol [27,28]), and perhaps even neurostimulation techniques (e.g., vagus nerve stimulation [29], and transcranial direct current stimulation [30]) can modify SDs in the brain. Therefore, detection and continuous monitoring of SDs as potentially treatable mechanisms of worsening brain damage may help improve outcome and pave the way towards precision health.

Brain injury is heterogeneous, which makes diagnosis and targeted therapy challenging [26,31,32]. Intracranial pressure (ICP) monitoring remains the cornerstone of neurocritical care for patients with severe TBI, yet it is an incomplete surrogate for the tissue-level pathophysiology driving secondary injury. Cerebral microdialysis studies have shown that metabolic crisis, marked by an elevated lactate/pyruvate ratio, persists in pericontusional tissue after TBI independent of cerebral perfusion pressure [33]. Multimodal monitoring cohorts have reinforced this discordance, finding that brain hypoxia and metabolic crisis are common in patients with acute brain injury despite a normal ICP [34]. SDs have emerged as a key driver of this hidden metabolic burden, with excitotoxicity and metabolic crisis accompanying SD events even when standard intensive care unit (ICU) parameters appear unremarkable [35]. Building on this mechanistic link, a prospective multicenter study from the Co-Operative Study on Brain Injury Depolarizations (COSBID) has demonstrated that SDs occur in over half of patients undergoing neurosurgery for TBI and are independently associated with unfavorable outcome after accounting for conventional prognostic covariates [36]. A subsequent, larger COSBID cohort confirmed that repetitive SDs, or clusters, are independently associated with poor neurologic recovery at six months, even when adjusted for other clinical and monitoring variables [19]. The DISCHARGE-1 study found that SD variables, in contrast to ICP, contributed to every multiple regression model of early, delayed, and total brain damage, patient outcome and death after SAH, strongly suggesting that SDs are also an independent biomarker of progressive brain injury in this disease [17]. Together, these findings indicate that ICP and other ICU measures alone are insufficient to capture the trajectory of secondary brain injury, motivating detection of SDs as a complementary monitoring modality that can provide earlier and more direct insight into ongoing cortical dysfunction.

### There is a need for noninvasive monitoring of SDs

Currently, gold-standard detection of SDs in clinical practice requires placement of subdural electrode strips during neurosurgical procedures. However, this monitoring method is inherently invasive, offers limited spatial coverage of the brain, and is available only to the small subset of patients who undergo invasive surgery. This subset represents a small fraction of the overall acute brain injury population: in the United States alone, an estimated 2.8 million TBI-related emergency department visits, hospitalizations, and deaths occur annually [37], yet only about 10% of cases are classified as severe [38]. The vast majority of TBI, roughly 70-90%, is mild [39], and even among moderate-to-severe cases managed in the hospital, most are treated conservatively without a craniotomy or decompressive procedure. A major goal in the field has therefore been to develop methods for noninvasive SD detection that can be deployed broadly across the neurocritical care population, to include the much larger population of acute cases with an intact skull, such as mild to moderate brain injury, concussion, and stroke.

Scalp EEG is noninvasive, widely available in ICUs worldwide, and offers global cortical coverage, making it an attractive candidate for noninvasive SD detection. However, its physics work against this goal: strong volume conduction through the skull acts as a spatial low-pass filter, blurring focal cortical activity and yielding poor spatial resolution, particularly for sources within sulci or deep cortex [40]. This blurring is reduced after decompressive hemicraniectomy, but the resulting skull defect introduces its own confound, breach rhythm, a high-amplitude, sharply contoured artifact arising from the loss of the skull’s filtering effect [41]. Scalp recordings are further limited by low signal-to-noise ratio, slow baseline drift, and non-propagating depressions, making visual identification of SDs difficult and time-consuming. These same distributed, subtle patterns, however, may be more tractable for automated detection algorithms than for human reviewers. To date, no clinically validated algorithm exists for reliable, noninvasive SD detection.

Previous attempts to detect SDs using EEG have had limited success. Early feasibility work relied on visual inspection of scalp EEG, with mixed and sometimes contradictory results. Hartings et al. [42] reported amplitude depressions associated with 81% of intracranially detected SDs through visual inspection in severe TBI patients, and Drenckhahn et al. [43] reported similarly positive results in malignant stroke and SAH patients, both using invasive electrocorticography (ECoG) as ground truth. Bastany et al. [44] described brief direct-current (DC) deflections in the scalp EEG of patients with epilepsy that they attributed to SD; however, these events were not confirmed invasively, and their short duration (approximately 23 s on average) and morphology differ from the scalp correlates of SD previously characterized against concurrent ECoG [42,43]. By contrast, Hofmeijer et al. [45] monitored 18 stroke and 18 TBI patients with 21-electrode EEG and found no SDs on visual inspection; because these patients neither underwent craniotomy nor had invasive recordings, it remains unclear whether SDs were even present. In a commentary on this negative result, Hartings et al. [46] cited the absence of established scalp-EEG detection criteria, spatial low-pass filtering by the intact skull, and short review epochs as possible confounds; using longer epochs instead, they identified candidate SD-like patterns in a TBI patient, again without invasive confirmation. In a separate cohort, Sivakumar et al. [14] monitored 15 TBI and 20 aneurysmal SAH (aSAH) patients with invasive ECoG and found no continuous scalp EEG pattern reliably associated with invasively confirmed SDs. In parallel, simulation-based studies explored automated, noninvasive SD detection independent of visual inspection, including an early forward-modeling and detection framework [47], a scalable deep-learning tracker [48] and graph attention network [49] for simulated SDs, and finite-element simulations of concussive SDs used to optimize DC-EEG electrode spacing [50].

More recently, related automated approaches have been tested on real human data. Wu et al. [51] reframed single-channel SD detection as a spectrogram-imaging task, adding a spectral dimension to the feature set, though validated only on augmented data from two patients (12 SDs total) and, by design, forgoing the spatial dynamics essential to tracking a traveling wave like SD. Brown et al. [52] introduced a dedicated high-resolution DC-EEG array (CerebroPatch) and reported proof-of-concept detection of SD-associated DC shifts against invasive ECoG in neuro-ICU patients, though usable, artifact-free data were obtained from only one of fifteen enrolled subjects. We developed WAVEFRONT [47,53], an automated, explainable algorithm, and used it to establish, for the first time, the feasibility of noninvasive SD detection in a single-center cohort of 12 severe TBI patients with decompressive hemicraniectomy, using simultaneous scalp and intracranial recordings as ground truth. Collectively, these efforts illustrate steady but incremental progress; however, it remains uncertain whether noninvasive SD detection generalizes beyond single-center cohorts, let alone whether it is sufficiently reliable for clinical use.

Prior noninvasive detection efforts, including our own single-center report [53], have been limited to small, homogeneous, single-institution cohorts, to visual inspection, or to simulation, leaving open whether a noninvasive SD biomarker can generalize across the clinical and anatomical heterogeneity of real-world acute brain injury, or extend meaningfully to the much larger population of patients with an intact skull. Here, we address these gaps directly. We train and rigorously validate WAVEFRONT, our automated, explainable scalp-EEG biomarker of SD, on a multicenter cohort of 32 patients spanning three institutions, three etiologies (TBI, SAH, and malignant hemispheric stroke [MHS]), and both skull-present and skull-absent conditions, using more than 1,100 intracranially confirmed SDs as ground truth. Using an exhaustive leave-four-out cross-validation framework, we quantify sensitivity, specificity, spatial specificity relative to the ground-truth electrode strip, temporal synchrony with individual and clustered SD events, and the ability to noninvasively estimate cumulative SD burden from continuous EEG.

Critically, we then test whether this biomarker, without retraining, generalizes to the far larger population lacking invasive ground truth: applying pre-trained WAVEFRONT to an independent, multicenter cohort of 607 intact-skull cardiac arrest patients from the International Cardiac Arrest REsearch (I-CARE) consortium, we show that noninvasively estimated SD burden is associated with neurological outcome. Together, these results provide multicenter, out-of-sample validation of a quantifiable noninvasive SD biomarker, motivating further studies to extend SD monitoring beyond the invasively monitored minority to the broader intact-skull population previously inaccessible to it.

## Methods

We first provide some details on the multi-center cohort used in this study, which includes 32 patients with mild to severe acquired brain injuries who underwent craniectomy or craniotomy (17 skull-present, 15 skull-absent patients) and intracranial ECoG strip electrode placement. We then describe the intracranial SD ground truth labeling, and introduce our automated SD detection method, emphasizing the explainability of WAVEFRONT by providing intuition for its main steps. Finally, we describe the external validation cohort of patients after cardiac arrest.

### Dataset Overview

The study dataset comprised 32 patients drawn from three neuromonitoring centers (referred to as Centers A, B, and C). These patients underwent simultaneous invasive ECoG and noninvasive scalp EEG recording. Across centers, recordings captured diverse pathologies, including aSAH, TBI, MHS, and acute subdural or epidural hematomas. Demographic and clinical characteristics, including age, sex, diagnosis, electrode location, skull condition, and the total number of spreading depolarizations (SDs), are included in Tables I–III.

**Table I.**
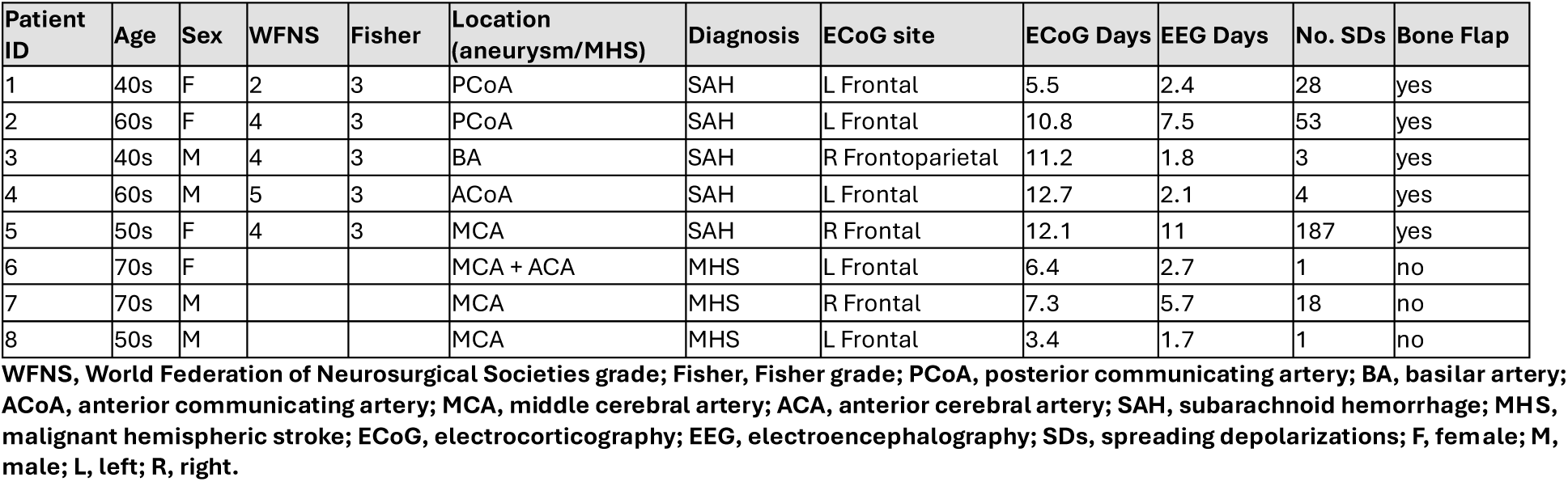
Clinical details and demographic information for five skull-present SAH and three skull-absent MHS patients in Center A.

| Patient ID | Age | Sex | WFNS | Fisher | Location (aneurysm/MHS) | Diagnosis | ECoG site | ECoG Days | EEG Days | No. SDs | Bone Flap |
| --- | --- | --- | --- | --- | --- | --- | --- | --- | --- | --- | --- |
| 1 | 40s | F | 2 | 3 | PCoA | SAH | L Frontal | 5.5 | 2.4 | 28 | yes |
| 2 | 60s | F | 4 | 3 | PCoA | SAH | L Frontal | 10.8 | 7.5 | 53 | yes |
| 3 | 40s | M | 4 | 3 | BA | SAH | R Frontoparietal | 11.2 | 1.8 | 3 | yes |
| 4 | 60s | M | 5 | 3 | ACoA | SAH | L Frontal | 12.7 | 2.1 | 4 | yes |
| 5 | 50s | F | 4 | 3 | MCA | SAH | R Frontal | 12.1 | 11 | 187 | yes |
| 6 | 70s | F |  |  | MCA + ACA | MHS | L Frontal | 6.4 | 2.7 | 1 | no |
| 7 | 70s | M |  |  | MCA | MHS | R Frontal | 7.3 | 5.7 | 18 | no |
| 8 | 50s | M |  |  | MCA | MHS | L Frontal | 3.4 | 1.7 | 1 | no |

WFNS, World Federation of Neurosurgical Societies grade; Fisher, Fisher grade; PCoA, posterior communicating artery; BA, basilar artery; ACoA, anterior communicating artery; MCA, middle cerebral artery; ACA, anterior cerebral artery; SAH, subarachnoid hemorrhage; MHS, malignant hemispheric stroke; ECoG, electrocorticography; EEG, electroencephalography; SDs, spreading depolarizations; F, female; M, male; L, left; R, right.

**Table II.** Clinical details and demographic information for 12 skull-present SAH and TBI patients in Center B.

| Patient ID | Age | Sex | HH | mFS | GCS | Diagnosis | ECoG site | ECoG Days | EEG Days | No. SDs | Bone Flap |
| --- | --- | --- | --- | --- | --- | --- | --- | --- | --- | --- | --- |
| 1 | 50s | F | 4 | 4 |  | aSAH | L Frontal | 9.8 | 12.3 | 0 | yes |
| 2 | 60s | M | 1 | 3 |  | aSAH | R Frontal | 8.9 | 8.9 | 0 | yes |
| 3 | 50s | M | 3 | 4 |  | mSAH | L Frontal | 4.8 | 2 | 27 | yes |
| 4 | 60s | F | 5 | 4 |  | aSAH | L Temporal + R Frontal | 3.5 | 4 | 61 | yes |
| 5 | 50s | F | 5 | 4 |  | aSAH | R Frontal | 6.5 | 8 | 2 | yes |
| 6 | 50s | F | 4 | 3 |  | aSAH | R Temporal | 8.9 | 6.3 | 4 | yes |
| 7 | 60s | M | 4 | 4 |  | aSAH | R Frontal | 11 | 12.2 | 24 | yes |
| 8 | 80s | M |  |  | 6 | TBI | R Frontal | 3.8 | 3.8 | 0 | yes |
| 9 | 20s | M |  |  | 5 | TBI | L Frontal | 5.7 | 10 | 0 | yes |
| 10 | 60s | F |  |  | 9 | TBI | L Frontal | 1.7 | 4.3 | 6 | yes |
| 11 | 40s | M |  |  | 14 | TBI | R Frontal | 10 | 9.8 | 0 | yes |
| 12 | 70s | F |  |  | 7 | SDH | R Frontal | 1 | 1 | 0 | yes |

HH, Hunt and Hess grade; mFS, modified Fisher scale; GCS, Glasgow Coma Scale; aSAH, aneurysmal subarachnoid hemorrhage; TBI, traumatic brain injury; SDH, subdural hematoma; other abbreviations as in Table I.

**Table III.** Clinical details and demographic information for 12 skull-absent TBI patients in Center C.

| Patient ID | Age | Sex | GCS | Pupils | Diagnosis | Lesion location | ECoG site | EEG/ECoG Days | No. SDs | Bone Flap |
| --- | --- | --- | --- | --- | --- | --- | --- | --- | --- | --- |
| 1 | 30s | M | 3 | 2 | TBI | R SDH | R Frontal | 3.1 | 4 | No |
| 2 | 40s | M | 3 | 0 | TBI | L SDH | L Frontal | 1.8 | 115 | No |
| 3 | 20s | M | 5T | 2 | TBI | R Cont, SDH | R Temporal | 4.7 | 128 | No |
| 4 | 20s | M | 2T | N/A | TBI | R A SDH | R Temporal | 3.9 | 181 | No |
| 5 | 70s | F | 2T | 2 | TBI | R A SDH | R Temporal | 0.3 | 71 | No |
| 6 | 70s | M | 14 | N/A | TBI | R SDH | R Temporal | 3.5 | 10 | No |
| 7 | 70s | M | 14 | 2 | TBI | R SDH | R Frontal | 3.6 | 0 | No |
| 8 | 60s | F | 7T | 2 | TBI | R SDH | R Parietal | 2.9 | 10 | No |
| 9 | 70s | M | 5T | N/A | TBI | R SDH | R Parietal | 5.5 | 57 | No |
| 10 | 40s | F | 6 | 2 | TBI | R SDH | R Frontal | 12 | 5 | No |
| 11 | 30s | M | 9 | 2 | TBI | R EDH | R Frontal | 0.5 | 0 | No |
| 12 | 20s | M | 3T | 2 | TBI | L SDH | L Temporal | 5.3 | 119 | No |

GCS, Glasgow Coma Scale (T, intubated); SDH, subdural hematoma; A SDH, acute subdural hematoma; Cont, contusion; EDH, epidural hematoma; N/A, not available; other abbreviations as in Tables I and II.

In Center C (ClinicalTrials.gov ID: NCT00803036), continuous EEG (cEEG) was recorded (95±42.2 hours on average) following craniectomy using a DC-coupled Moberg amplifier (CNS Advanced ICU EEG Amplifier from MOBERG ICU Solutions), with standard ICU montage of 19 individually placed electrodes on scalp and a sampling frequency of 256Hz. ECoG was recorded using the same amplifier. In Center B, ECoG recordings were acquired using DC-coupled Moberg amplifier and used for SD ground truth annotations (see *ECoG SD annotations*), while cEEG was recorded using either the same amplifier (four TBI patients), or separate amplifier (Natus Xltek Brain Monitor, one TBI and seven SAH patients). For these eight patients, we synchronized the cEEG and ECoG amplifiers using the common High-Frequency (HF)-ECoG (>0.5Hz) channels recorded using both amplifiers. This was done by power-envelope extraction and cross-correlation of a representative channel segment with good recording quality, identified through visual inspection of the ECoG signals. This amplifier synchronization technique enables utilization of a larger cohort of ICU data for non-invasive SD detection studies in the future. In Center A, ECoG data was recorded using GT205 amplifier using Powerlab A/D converter (ADInstruments), while scalp EEG was recorded using DC-coupled BrainAmp (BrainAmp MR plus, Brain Products). ECoG and scalp EEG recording started on separate laptops with manual synchronization (person starting the recording had to press a key on both systems simultaneously). Thus, the synchronization is not perfect. However, since SD is a slow event, with the speed of mm/min, a synchronization error of a few seconds or less is adequate for the SD detection task, where the temporal resolution for calculation of performance metrics is in the order of minutes (see Results).

#### ECoG SD annotations

In all centers, during the craniectomy/craniotomy procedure, or through a burr hole in the skull, a strip of six monopolar ECoG electrodes, with an interelectrode distance of 1cm, was placed on the ipsilateral hemisphere, close to the site of injury. The recorded ECoG signals were visually assessed by clinical experts to identify and annotate the SD episodes in the dataset.

SDs were identified and annotated following recommendations by the Co-Operative Studies on Brain Injury Depolarizations (COSBID): In this study, an episode of SD refers to a unique SD propagation, while considering all electrodes of the ECoG strip. For each episode, the starting time point of SD was annotated based on a chosen ECoG electrode, which is not necessarily the same across episodes/patients. This temporal annotation may not reflect the actual starting time point of the SD wave because of (i) the limited spatial coverage of a single ECoG strip, as SDs may originate outside this coverage, and (ii) inherent variability in visual inspection of the episodes. Limited strip coverage also implies that some SDs are not captured by ECoG at all: many SDs in the human cortex are spatially restricted[5], whereas scalp EEG samples a much larger cortical area. Because detection performance was scored in time windows against strip annotations (see Results), any WAVEFRONT detection without a concurrent ECoG-annotated SD was counted as a false positive, regardless of where on the scalp it occurred. Some of these apparent false alarms may therefore correspond to genuine SDs outside the strip coverage, in which case the reported false positive rates overestimate the true false-alarm rate.

#### Demographics

The cohort included 19 males and 13 females, with an average age of 55.2±16.4 years, reflecting the typical age span of neurocritical care populations monitored for SD activity. Mean age was 59.8, 58.9, and 48.5 years in Centers A, B, and C, respectively, with Center A contributing primarily aneurysmal SAH cases, Center B predominantly additional SAH patients, and Center C consisting entirely of TBI patients, predominantly with acute subdural hematomas.

#### Pathology and Etiology Distribution

The cohort exhibited a broad distribution of neurological etiologies associated with susceptibility to spreading depolarizations. Aneurysmal and non-aneurysmal subarachnoid hemorrhage (aSAH, SAH, mSAH) represented a substantial portion of the dataset, with additional cases of TBI, MHS, and acute subdural or epidural hematomas. Several patients presented with mixed injury patterns, including combinations of contusions and extra-axial hemorrhage. This range of pathologies reflects the typical spectrum of disorders encountered in neurocritical care settings where SD monitoring is clinically relevant.

#### Electrode Location and neurosurgical approach

ECoG strip placement varied according to the neurosurgical approach, including frontal, temporal, parietal, or combined exposures. Neurosurgical approach and therefore skull condition (bone flap present/replaced vs absent) differed systematically among centers; TBI patients in Center C and MHS patients in Center A underwent craniectomy, whereas all other patients underwent craniotomy. EEG electrode montages followed standardized 10–20-based coverage in Center B and C, while partial montage was used in Center A with more ipsilateral coverage. In the invasive-monitoring cohort, we only use a subset of scalp electrodes in the ipsilateral hemisphere and therefore this montage variation across centers is unlikely to affect the results.

#### SD Burden

A total of 1,119 SDs were annotated on ECoG across the 32 patients. The SD burden varies widely across subjects, with some demonstrating minimal to zero SD events and others exhibiting prolonged recurrent SDs (up to 187 episodes) over extended monitoring durations. This heterogeneity is consistent with prior ECoG-based SD studies [54].

### Preprocessing pipeline

As noted in Dataset Overview, EEG recordings in this cohort originate from two different acquisition systems: the CNS Advanced ICU EEG Amplifier (MOBERG ICU Solutions), used with or without continuous electrode-impedance logging, and the Natus Xltek system. Because these two setups differ in what electrode-level quality information is available, we adapted a multi-step preprocessing pipeline, built on the framework we introduced in [53], separately to each recording condition and data format, as detailed below.

#### Band-pass filtering and downsampling

Our earlier work [53], consistent with independent findings in the field [42], identified the Delta band (0.5–4 Hz) as the frequency range yielding the strongest non-invasive SD detection performance. Guided by this, scalp EEG signals here were band-pass filtered to isolate the Delta band using a Hamming-windowed sinc finite impulse response (FIR) filter (filter order 1000; transition bandwidth 0.02 Hz), then downsampled to 10 Hz, comfortably above the Nyquist rate for this band, to reduce the computational load of the downstream analysis.

#### Identifying and excluding poor-quality segments of EEG signal

Continuous ICU recordings inevitably contain stretches degraded by environmental and procedural noise, for instance amplifier disconnection while a patient is moved for imaging or a bathroom break, or transient loss of electrode-scalp contact from patient movement. For the 12 patients whose recordings included continuous impedance logging (CNS MOBERG system), we adapted the impedance-based masking approach from [53]: at each electrode, intervals whose recorded impedance exceeded twice the electrode’s own median impedance (2M_ch_) across the full recording were flagged and excluded from the subsequent SD-detection steps. For the remaining 20 patients, no impedance channel was available, so poor-quality segments were identified solely through the amplitude-outlier step described next. Retaining both types of recordings lets us test WAVEFRONT’s robustness under the data-quality conditions most ICUs actually have (typically without impedance monitoring), rather than only under best-case conditions.

#### Outlier removal

The final step of preprocessing addresses residual amplitude artifacts. We flagged abnormally large positive or negative excursions using Tukey’s fences [55]: a sample is treated as an outlier if it falls below Q_1_ − k(Q_3_ − Q_1_) or above Q_3_ + k(Q_3_ − Q_1_), where Q_1_ and Q_3_ are the signal’s first and third quartiles and k sets the stringency of the criterion. We used k = 3, the conventional threshold for ‘far-out’ extreme values, and masked the corresponding segments at each electrode. Supplementary Fig. 2 in [53] illustrates this preprocessing sequence applied to ipsilateral EEG from a representative skull-absent TBI patient over a four-hour window. The cleaned signals were then passed to WAVEFRONT for SD detection.

### Noninvasive WAVEFRONT biomarker detection on scalp

We applied an enhanced version of the WAVEFRONT algorithm, originally introduced in [47] and clinically validated in [53], to detect and characterize noninvasive SD signatures, which we refer to as WAVEFRONT biomarkers, in scalp EEG from all 32 patients in this cohort (Tables I–III). WAVEFRONT is built to be explainable: every intermediate representation and the final detection output are interpretable, in contrast to end-to-end deep learning approaches whose internal decision process is not explainable. Rather than learning SD signatures purely from data, WAVEFRONT’s architecture is constructed around established physical properties of SD waves (their propagation speed, the spatial width of the associated suppression, and their characteristic spread pattern), so that the detection logic reflects known SD biophysics rather than an unconstrained pattern-matching process. This design directly targets the core difficulty of scalp-based detection: SD signals arriving at the scalp are weak, noisy, and spatially blurred by the intervening tissue. WAVEFRONT addresses this by breaking the detection problem into a sequence of smaller, more tractable subproblems, each aimed at isolating and classifying individual SD wavefronts rather than attempting to detect the whole propagating event at once. Concretely, power envelopes are first computed from each electrode’s filtered signal, and intervals of sustained power suppression are extracted; these suppressed intervals are then mapped onto a two-dimensional representation of the electrode array, producing discrete candidate wavefronts. WAVEFRONT then tests each candidate for genuine propagation by estimating its velocity vector using optical flow, a standard computer-vision technique that infers displacement from frame-to-frame brightness changes; here, the ‘frames’ are successive snapshots of the spatial suppression maps rather than conventional video. Linking these local propagation estimates across time and space lets the algorithm reconstruct the larger-scale trajectory of SD activity across the cortex, a strategy that also helps absorb distortions introduced by cortical folding (sulci and gyri) and supports detection of more complex, multidirectional spread patterns.

We closely follow the steps of the WAVEFRONT algorithm, proposed in [53]:

1. *Epoching, Envelope Extraction, and Depression Identification:* Cleaned EEG recordings are divided into fixed-length epochs, and a power envelope is computed for each electrode’s band-limited signal within each epoch. Within these envelopes, we flag depression events (transient drops in amplitude consistent with an SD) using a correlation-based detector tuned to recognize the characteristic first-derivative-like decline at the onset of a depression. The result is a time-resolved catalog of candidate depressions across the electrode array, which feeds the spatial reconstruction step that follows.
2. *Spatial Projection and Reconstruction of Depression Wavefronts:* Candidate depression events, initially defined on the three-dimensional scalp electrode layout, are mapped onto a two-dimensional cylindrical projection of the scalp. The resulting sparse spatial map is interpolated, thresholded, and downsampled into a binary image in which connected depressed regions represent candidate wavefronts. Working in this flattened 2D projection removes distortions caused by the irregular electrode geometry and makes the subsequent characterization of spatial spread more tractable.
3. *Optical Flow–Based Motion Estimation:* For each pair of consecutive binary images, WAVEFRONT computes a dense optical-flow field to characterize how candidate wavefronts move between frames. The resulting local velocity vectors are pooled into spatially contiguous groups (‘bounding boxes’, or BBoxes), and a directional histogram is built for each BBox to identify its dominant direction(s) of motion. This pooling step is what lets the algorithm recover a stable, mesoscale estimate of propagation direction while averaging out pixel-level noise in the flow field.
4. *Orientation Bounding Boxes, Effective Propagation Measure, and Consistency Scoring*: BBoxes sharing a common dominant orientation are merged into Orientation Bounding Boxes (OBBoxes). Before further scoring, each OBBox is tested against the Effective Propagation Measure (EPM), a metric that separates genuine translational movement from static fluctuations or artifactual ‘pop-up’/‘fade’ patterns that mimic movement without true spatial spread. OBBoxes falling below a learned EPM threshold are discarded. In this study, we revised the original EPM formulation of [53] as follows:

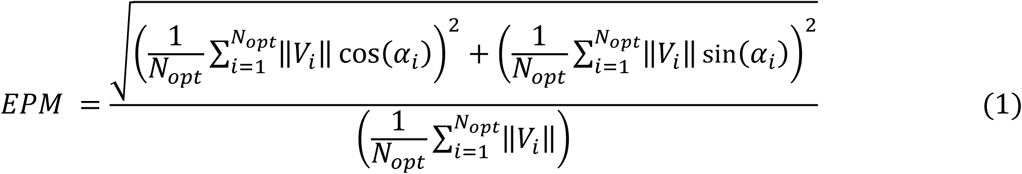

where ‖*V_i_*‖ and α_i_ are, respectively, the magnitude and orientation of the i-th optical-flow vector within the OBBox, and *N_opt_* is the total number of flow vectors it contains. We normalize the resultant vector by the average flow magnitude in the box; this rescales EPM to a fixed [0, 1] range, with 0 indicating no net translational movement and 1 indicating fully coherent, unidirectional propagation, which in turn makes it possible to learn a single generalizable EPM threshold across boxes of different overall motion magnitude (see Results). OBBoxes that pass the EPM test then undergo spatiotemporal consistency scoring against their neighbors, checked for agreement in direction, speed, and timing, and candidates whose implied propagation speed falls outside physiologically plausible bounds are excluded, so that only biologically credible propagation patterns are carried forward.
5. *Frame Selection, Temporal Stitching, and Final SD Detection* Frames whose surviving OBBoxes do not meet minimum coherence criteria are dropped, based on frame-level scores derived from the underlying OBBox statistics. The remaining positive frames are then linked across time with a sliding temporal window to form a continuous detection trace. A given frame is ultimately labeled as containing an SD when it shows sustained, directionally coherent, spatially contiguous wavefront propagation supported by OBBoxes that have passed both the EPM and consistency checks. The output of this step is a time-resolved trace of SD detections that can be directly compared against the intracranial ground truth (see Results).

### External validation cohort (I-CARE)

To assess the biomarker in patients with an intact skull, we analyzed continuous EEG from the 607 patients in the publicly released portion of the multicenter I-CARE database [56], which includes comatose patients aged 16 years or older who underwent continuous EEG monitoring after return of spontaneous circulation following cardiac arrest. No invasive recordings are available in this cohort. The WAVEFRONT model trained on the full 32-patient invasive-monitoring cohort was applied without retraining to the full bilateral montage of 19 scalp electrodes at standard 10–20 locations, so that SD burden reflects detections across the whole scalp. SD burden was defined as the percentage of time with WAVEFRONT-detected SD; it was computed in 12-h windows advanced in 1-h steps, with bootstrap 95% confidence intervals, and summarized over consecutive 24-h periods measured from the start of each EEG recording (0–24, 24–48, and 48–72 h) and the cumulative 0–72 h period. Neurological outcome was assessed with the Cerebral Performance Category (CPC) scale and classified as good (CPC 1–2) or poor (CPC 3–4). Patients who died (CPC 5) were excluded from outcome comparisons because death after cardiac arrest may reflect decisions to withdraw life-sustaining treatment in addition to neurological injury. Group differences in SD burden were summarized with Cohen’s d, and odds ratios (OR) for poor outcome were expressed per 10% increase in SD burden.

## Results

We validate the performance of our WAVEFRONT technique on this multicenter cohort across three centers, three neurological conditions (TBI, SAH, and MHS), and two neurosurgical procedures (craniectomy or skull-absent, and craniotomy or skull-present). Intracranial recordings identified 1,119 SDs, providing a unique dataset with broad representation of SD morphology, frequency, and recurrence patterns. This heterogeneous SD dataset allows for within- and out-of-sample validations, assessing the spatiotemporal specificity of the detected events, and quantification of SD burden in long time intervals.

### Cross-Validation performance across all patients

Algorithmic performance is quantified using a leave-four-out cross-validation framework: in each fold, four patients are held out for validation, and the remaining 28 patients are used to train WAVEFRONT. We perform exhaustive cross-validation over all possible train-validation sets and obtain average receiver-operating characteristic (ROC) curves (Fig. 1). The optimal operating point is found by minimizing a validation error which is defined as the distance to the ideal operating point (true positive rate [TPR] = 1; false positive rate [FPR] = 0), i.e., *ε_CV_* = √*FPR*^2^ + (1 − *TPR*)^2^. This cross-validation error *ε_CV_* provides a tradeoff between low false-alarm rate and high detection rate. The resulting optimal operating point is a “knee” point on the validation curve (Fig. 1, right inset). WAVEFRONT achieved a true positive rate of 0.973 ± 0.01 and a false positive rate of 0.060 ± 0.005 (95% CI). These performance metrics are defined and calculated over 2-min sliding windows with 30-s overlaps, closely following the steps in [53].

**Figure 1.**
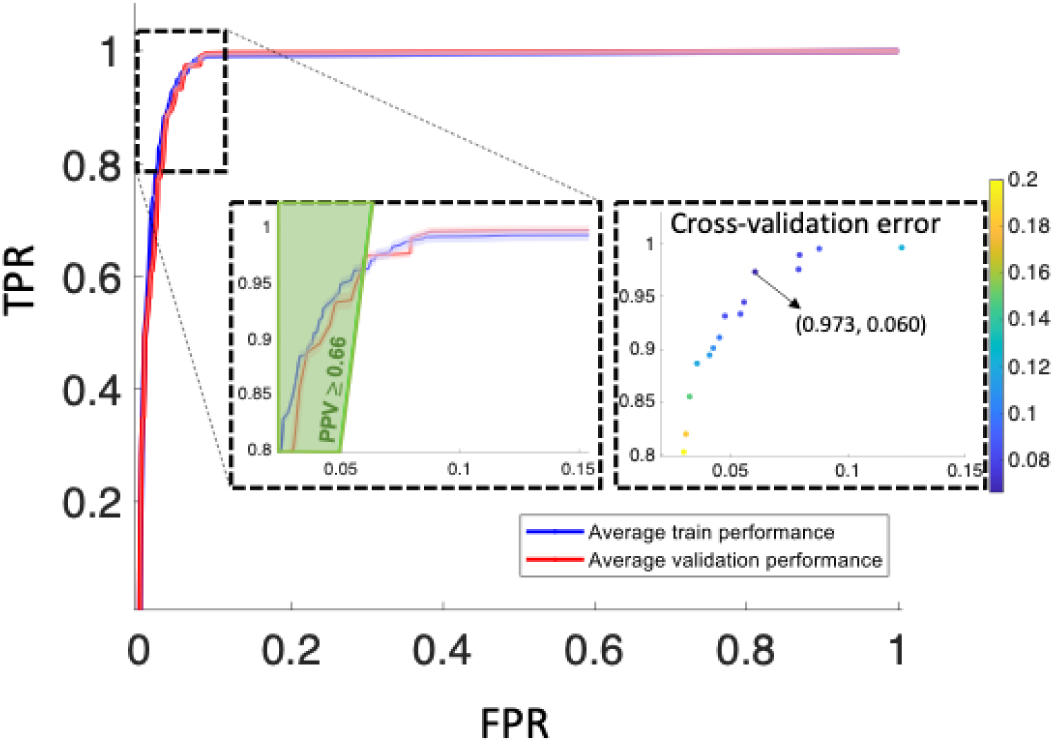
WAVEFRONT performance evaluation on all 32 patients using a cross-validation analysis. Receiver operating characteristic (ROC) curves of the average train (solid blue curve) and validation (solid red curve) performance of WAVEFRONT in the detection of spreading depolarization (SD) events, using noninvasive scalp electroencephalography (EEG) signals in the Delta frequency band. The shaded blue and red regions show the 95% confidence intervals for the average train and validation curves, respectively. The vertical axis indicates true positive rates (TPRs), and the horizontal axis shows false positive rates (FPRs). The highlighted green region indicates the positive predictive values of ≥0.66 (PPV). The left inset is the zoomed-in version of the ROC curves around the optimal validation operating point (TPR= 0.973±0.01, and FPR= 0.060 ±0.005; in 95% confidence intervals). The right inset is the cross-validation error, color-coded across different points in the validation ROC curve, where the point with the minimum error (i.e., optimal operating point) is marked.

The positive predictive value (PPV) surface (Fig. 1 left inset, shaded green region) exhibits a wide region where PPV ≥ 0.66, i.e., for a wide range of operating points on the validation curve, more than 66% of WAVEFRONT detections correspond to ECoG-confirmed SDs.

### Spatial specificity of scalp-derived SD signatures

Spatial consistency of WAVEFRONT biomarkers with the ground truth SDs is assessed by visualization of detected power depression on scalp topography, where the location of invasive strip is marked (Fig. 2). For example, in a skull-absent patient with TBI (Fig. 2, top row), the WAVEFRONT-detected depression propagated over the right frontotemporal region (Fig. 2b, right panel), where the ECoG strip was located and recorded a propagating SD (Fig. 2b, left panel). In another example, for a skull-present TBI patient (Fig. 2, bottom row), a similar spatial specificity was observed: despite the presence of the low-conductive skull in the perilesional hemisphere during the recording, scalp-detected depressions were spatially consistent with the location of the ECoG strip in the left occipitotemporal lobe (Fig. 2b, right panel). We also observed that this spatial consistency is unique to the cases where there is a propagating SD, i.e., in recordings without any observed/annotated SD event, no WAVEFRONT propagating depressions were observed around the strip location. A 1-hour no-SD recording interval is shown in Fig. 2b for a skull-present SAH patient (middle row), where no coherent propagating depression was observed around the ECoG strip.

**Figure 2.**
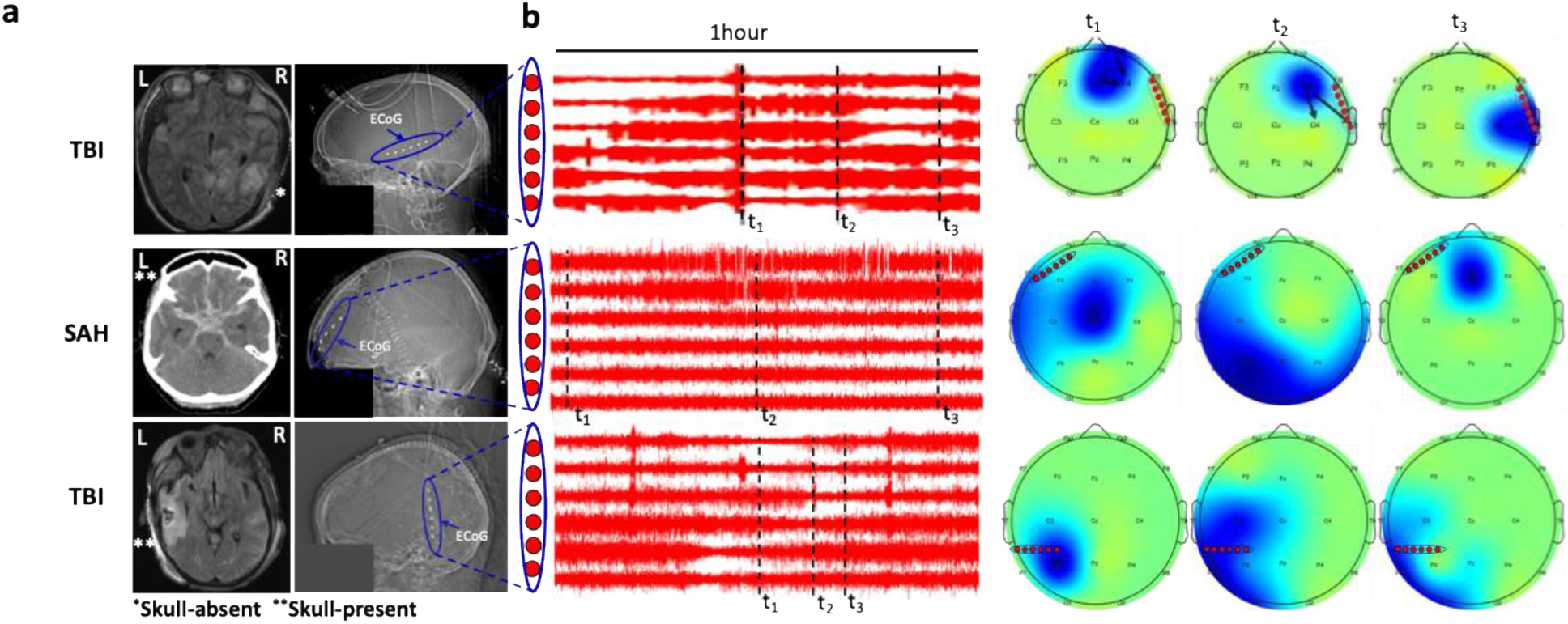
Spatial specificity of scalp-detected SD signatures in brain injury patients. (a) Transverse and side view of computed tomography (CT) and magnetic resonance imaging (MRI) scans of a skull-absent TBI patient with right temporal strip at right decompressive hemicraniectomy (DHC), and two skull-present patients with replaced bone flaps (a SAH with left frontal strip, a TBI with left temporal strip). The missing portion of the skull (marked with **), the replaced bone flap (marked with *), and the strip of six electrocorticography (ECoG) electrodes are also marked. The facial region is stripped away for anonymity. (b-left panel) one-hour intervals across 6 ECoG electrodes for each of the three patients in (a), (b-right panel) scalp topography of detected power depressions (blue spots) in electroencephalography (EEG), for each of the three time points annotated in the ECoG one-hour intervals. For the two TBI patients, episodes with SD events are shown in (b), while a negative episode (i.e., no SD event) is shown for the SAH patient. SD power depression (blue spot) propagation is observed around the location of the strip (b, right panel), which shows the spatial consistency and specificity of scalp WAVEFRONT biomarkers with strip ground truth of SD. In the SAH patient without any ground truth SD, no consistent propagation was observed across the three time points in the 1-hour interval.

### Generalization Across Centers, Etiologies, and Skull Conditions

We evaluated algorithmic stability across clinical sites, diagnostic categories, and skull configurations. WAVEFRONT maintained consistent performance under within-sample and out-of-sample evaluations (Fig. 3a). When trained on patients from one center and tested on another, detection performance remained stable, demonstrating resilience to differences in acquisition hardware, ICU noise environments, and montage variations.

**Figure 3.**
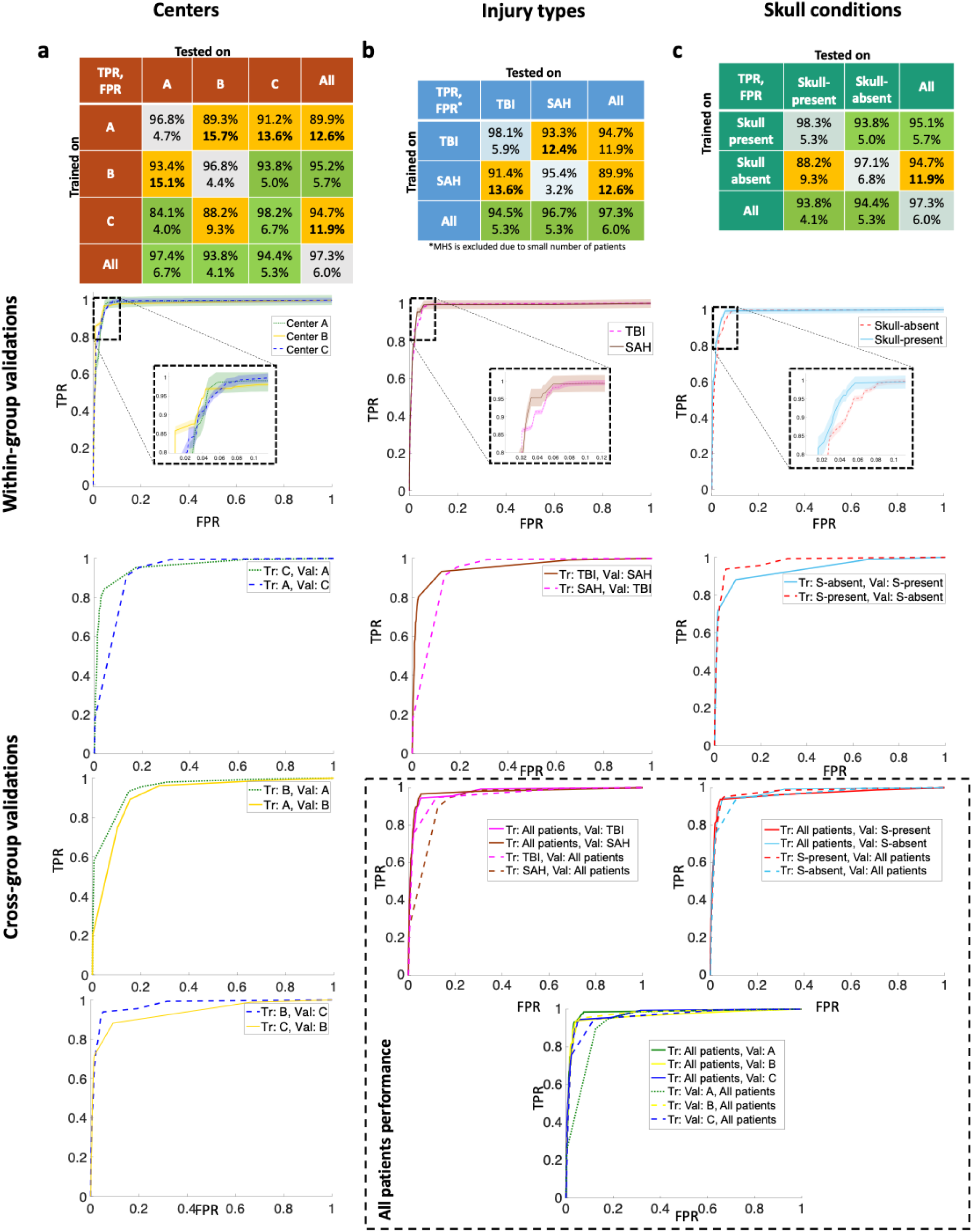
Within-sample and out-of-sample validations across centers, injuries, skull conditions. This figure summarizes the overall performance of WAVEFRONT algorithm under two scenarios: trained and validated on the same group (within-sample validation, diagonal values in the performance tables) and trained and validated on different groups (out-of-sample validation). MHS was excluded from the injury-type analysis because too few patients (n=3) were available for reliable performance estimation. The within-sample performance is obtained through leave-four-out exhaustive cross-validation, averaged over all validation sets. WAVEFRONT achieved strong within-sample SD detection performance across all groups, i.e., three centers, two injury types, and skull conditions with >95% TPR and <7% FPR. Based on the within-sample validation results, WAVEFRONT, once trained on large enough data, performs well on each of the patient groups. The out-of-sample validation performance is obtained by training the model on one group, e.g., Center A, finding the optimal set of thresholds (*Thr*_1_*^opt^*, *Thr*_2_*^opt^*), and applying the trained WAVEFRONT on the other group, e.g., Center B. Validation curves are obtained across a predefined range of thresholds *Thr*_3_*^opt^*, *Thr*_4_*^opt^*). Strong generalizability was observed from skull-present to skull-absent patients (93.8% TPR, 5.0% FPR out-of-sample average performance). More importantly, the out-of-sample validation results show that WAVEFRONT, once trained on all groups of patients with different conditions, performs well on each of the subgroups.

Stratification by etiology (TBI vs. SAH; MHS was not analyzed separately because only three patients were available) showed comparable performance (Fig. 3b). Similarly, performance was preserved in both skull-present and skull-absent patients (Fig. 3c), indicating that major anatomical differences in cranial filtering did not degrade detection quality. These results indicate that WAVEFRONT captures cortical depression dynamics that generalize across key clinical, anatomical, and technical sources of variability.

### Temporal Synchrony Between Scalp and Intracranial SDs

We examined temporal alignment between scalp-derived depression detections and intracranial SDs using representative examples (Fig. 4b). WAVEFRONT detections showed close temporal correspondence with the onset, evolution, and recovery phases of SDs recorded on ECoG. This synchrony was preserved across centers, across skull conditions, and across etiologies.

**Figure 4.**
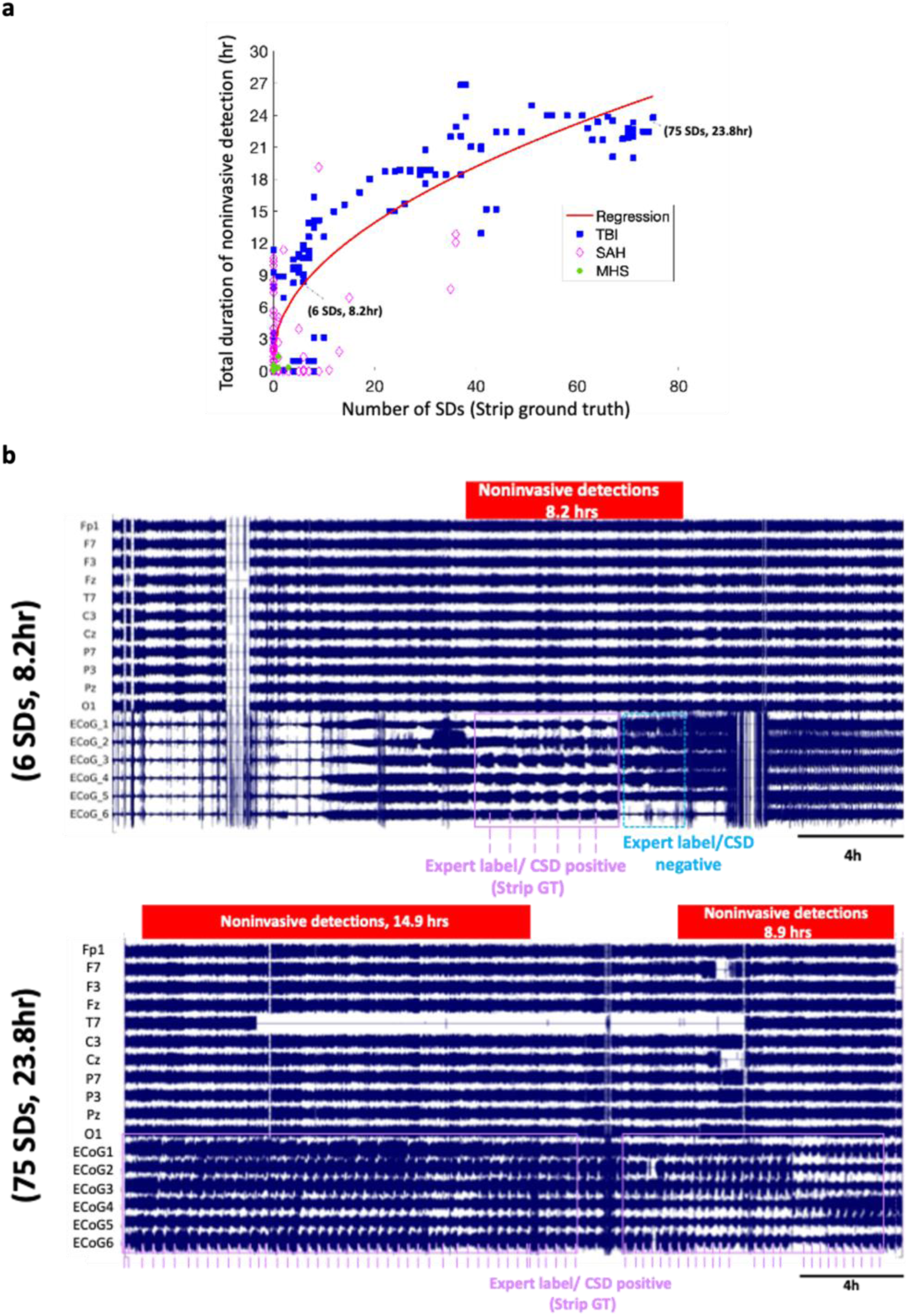
Quantification of WAVEFRONT biomarker time-burden and visualization of its temporal synchrony in continuous EEG vs ECoG. (a) Estimation of number of SDs in 30-hour time windows (horizontal axis) using WAVEFRONT duration of detection (vertical axis). The biomarker alarm time-burden is correlated with the frequency of occurrence of SDs (R^2^=0.7411), (b) temporal synchrony via visual inspection.

During clustered SD activity, scalp EEG depressions tracked prolonged suppression periods corresponding to multiple, closely spaced SDs, while occasional under-segmentation occurred only during the densest clusters, consistent with known challenges even in invasive recordings (Fig. 4b).

### Quantification of SD Burden From Continuous EEG

To determine whether WAVEFRONT could provide a quantitative measure of SD burden, we compared the total duration of scalp-detected depression episodes with the number of ECoG-confirmed SDs in overlapping 30-hour windows (Fig. 4a). The resulting relationship was strongly linear, yielding an R² of 0.7411, demonstrating that cumulative depression duration reflects underlying SD load. Because SD burden correlates with metabolic crisis and lesion progression, the ability to estimate this measure noninvasively from standard EEG may represent a clinically meaningful advance for continuous neuromonitoring.

WAVEFRONT achieved robust noninvasive SD detection using a stringent leave-four-out cross-validation framework, demonstrating consistent performance on entirely unseen patients. The method showed close spatial correspondence between scalp-derived depressions and invasive ground truth in representative skull-present and skull-absent cases and exhibited strong generalization across centers and etiologies indicating stability across diverse clinical environments. Scalp detections maintained high temporal fidelity with individual SDs, accurately tracking both isolated and clustered events. Finally, WAVEFRONT provided reliable quantitative estimation of SD burden over long ICU recordings, paving the way towards noninvasive measurement of cumulative SD activity.

### External Validation of WAVEFRONT SD Burden After Cardiac Arrest

All analyses above were performed in neurosurgical patients with concurrent ECoG. We next asked whether the biomarker carries clinically relevant information in patients with an intact skull, in whom SDs cannot be recorded invasively. To this end, the WAVEFRONT model trained on the 32-patient invasive-monitoring cohort was applied, without retraining, to continuous EEG from 607 patients who were comatose after cardiac arrest, drawn from the multicenter I-CARE consortium (see Methods). The primary comparison was between survivors with good (CPC 1–2, n=225) and poor (CPC 3–4, n=29) outcome. SD burden in the first 24 hours of post-arrest EEG recording did not differ between these groups (Cohen’s d = −0.012; OR 0.995; p=0.95). Separation emerged after the first day (Fig. 5): on day 2 (24–48 h), SD burden tended to be higher in the poor-outcome group (OR 1.106; p=0.068), and on day 3 (48–72 h) the difference was largest and statistically significant (Cohen’s d = 0.556; OR 1.151 per 10% increase; 95% CI 1.040–1.275; p=0.007). SD burden accumulated over the full 0–72 h window was likewise higher in the poor-outcome group (Cohen’s d = 0.457; p=0.024). Patients who died (CPC 5) had consistently lower SD burden throughout monitoring. Two factors may contribute to this finding. First, death in this population may reflect decisions to withdraw life-sustaining treatment, not only the severity of neurological injury. Second, WAVEFRONT detects depressions of ongoing activity, which cannot be resolved in maximally suppressed cortex; low burden in this group may therefore reflect a detection ceiling rather than a true absence of SDs. This consideration motivates including EEG suppression ratio as a covariate when SD burden is used for prognostication.

**Figure 5.**
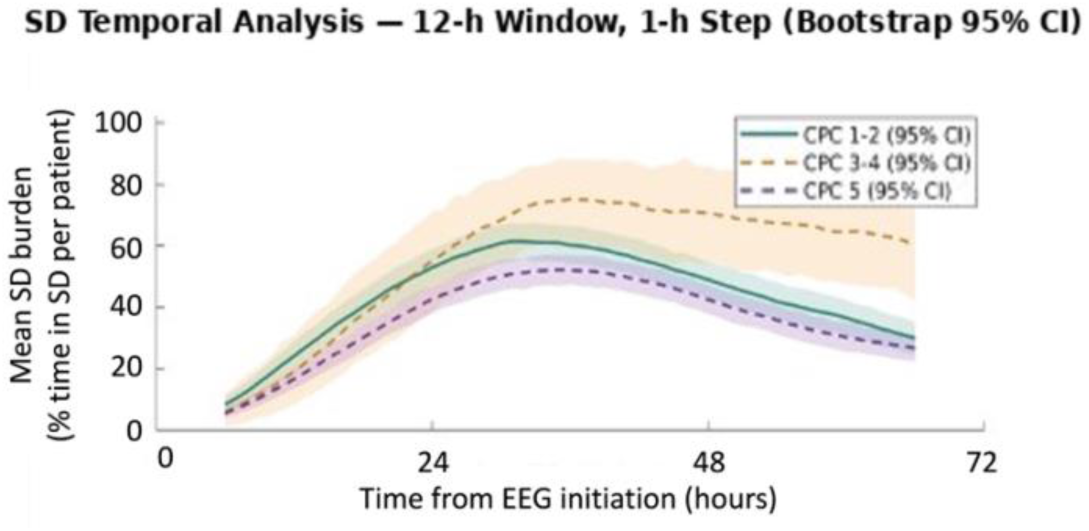
**External validation of** the pre-trained WAVEFRONT biomarker in intact-skull patients after cardiac arrest. Mean SD burden (percentage of time with WAVEFRONT-detected SD per patient), computed in 12-h windows advanced in 1-h steps, is shown for survivors with good (Cerebral Performance Category [CPC] 1–2) and poor (CPC 3–4) neurological outcome and for patients who died (CPC 5). Time is measured from the start of post-arrest EEG recording. Shaded areas indicate bootstrap 95% confidence intervals.

## Discussion

In a multicenter cohort spanning three institutions, three etiologies, and both skull-present and skull-absent conditions, we validated a quantifiable noninvasive biomarker of SD in out-of-sample analyses. Using more than 1,100 intracranially confirmed events as ground truth, WAVEFRONT achieved a true positive rate of 0.973 with a false positive rate of 0.060 under stringent leave-four-out cross-validation, with performance preserved across centers, diagnoses, and skull conditions, addressing the central limitation of prior noninvasive SD efforts, which were confined to single institutions, small or homogeneous cohorts, or simulation, and had not been tested for generalization across the clinical and anatomical heterogeneity of real-world neurocritical care.

Beyond detection accuracy, WAVEFRONT’s scalp-derived depressions were spatially consistent with the location of the underlying ECoG strip in representative cases and were absent during intervals without ground-truth SDs, arguing against the biomarker simply tracking generic cortical slowing or artifact. Its explainable, biophysics-constrained architecture, in which detection stages map onto known SD propagation speed, spatial width, and directional coherence, distinguishes it from end-to-end deep-learning approaches whose decision process is opaque, a property likely to matter for clinical adoption and regulatory scrutiny of a novel neuromonitoring biomarker. WAVEFRONT also provided a quantitative estimate of cumulative SD burden that correlated strongly with intracranially confirmed SD frequency, suggesting that continuous, noninvasive burden estimation, rather than binary event detection alone, may be clinically actionable given the established relationship between SD burden and secondary injury.

The extension to the I-CARE cardiac arrest cohort is, to our knowledge, the first demonstration that a scalp-EEG SD biomarker trained exclusively on invasively confirmed, surgically treated patients can be applied, without retraining, to a much larger population for whom invasive confirmation is never obtained. That SD burden on day 3 of post-arrest EEG monitoring was associated with neurological outcome in this entirely intact-skull cohort supports the premise motivating this work, that SD monitoring need not remain confined to the small, surgically selected minority of acute brain injury patients, but may generalize to the broader population, including mild-to-moderate TBI, stroke, and other intact-skull etiologies, for whom no other means of SD detection currently exists. The consistently lower SD burden observed in patients who ultimately died (CPC 5) may reflect withdrawal of life-sustaining treatment and a detection ceiling in maximally suppressed cortex rather than a true absence of SD activity, underscoring that suppression ratio should be considered alongside SD burden in future prognostic models.

Several limitations warrant consideration. Ground-truth SD annotation relied on a single ECoG strip with limited spatial coverage; SDs originating outside this coverage could be missed or misattributed, and, because many SDs are spatially restricted [5], some scalp detections counted as false alarms may reflect genuine SDs beyond the reach of the strip. Region-resolved analyses of detections remote from the strip, ideally with multiple strips as ground truth, will be needed to determine how many apparent false alarms represent true SDs. The training cohort, while larger and more heterogeneous than prior single-center efforts, remains modest in absolute size, and cross-institutional differences in amplifier hardware and montage, though shown not to degrade performance here, warrant confirmation in additional independent centers. Critically, the I-CARE validation could not be benchmarked against invasive ground truth by design, since intact-skull patients do not undergo ECoG monitoring; the outcome association we observe is therefore prognostic evidence of biological relevance rather than direct confirmation of detection accuracy in this population. Additionally, our training and validation cohorts consisted exclusively of adults, and I-CARE included only patients aged 16 years or older, so the biomarker’s performance in pediatric populations, who differ in skull thickness, cortical maturation, and SD susceptibility, remains untested.

A particularly timely application is SD-targeted pharmacotherapy. The ongoing INDICT trial (Improving Neurotrauma by Depolarization Inhibition with Combination Therapy), a phase 2 randomized feasibility trial led by Hartings and colleagues across three US centers [26], uses a tiered treatment protocol, including ketamine, to suppress SDs in patients with severe TBI, but by design requires surgical placement of a subdural strip electrode for both enrollment and treatment guidance, restricting the trial to patients who undergo neurosurgery. A validated noninvasive biomarker like WAVEFRONT could extend this paradigm in two ways: first, by enabling noninvasive screening or confirmation of SD activity to complement or reduce reliance on invasive monitoring within trials structured like INDICT, and second, and more broadly, by opening SD-guided ketamine titration to the far larger population of TBI, stroke, and other acute brain injury patients who never undergo strip placement and are therefore categorically excluded from current SD-targeted trials. Realizing this will require prospective studies directly comparing noninvasive and invasive SD burden as triggers for ketamine dosing; the present results provide a foundation for such studies.

Future work should also prioritize larger, prospective multicenter validation; extension to additional intact-skull etiologies such as mild TBI, migraine, and non-surgical stroke; and extension to pediatric cohorts, where noninvasive SD monitoring could be particularly valuable given the added ethical and practical barriers to invasive neuromonitoring in children.

## Data Availability

The dataset was obtained as part of a multicenter clinical study. The anonymized raw EEG dataset may be made available upon request, contingent upon contractual obligations and data sharing and cooperative agreements.

## Acknowledgements

This work was supported, in part, by grants from Chuck Noll Foundation for Brain Injury Research, the Office of the Assistant Secretary of Defense for Health Affairs through the Defense Medical Research and Development Program under Award No. W81XWH-16-2-0020, Hillman Foundation, and a P30 CORE award EY08098 from the National Eye Institute, NIH. A Chamanzar was also supported by the Roche Postdoctoral Fellowship at CMU Neuroscience Institute. JP Dreier was supported by DFG DR 323/10-2 (413848220), DFG DR 323/13-1 (570363796), EU Horizon MSCA-DN 101119916 – SOPRANI and HORIZON-EIC-2026-PATHFINDEROPEN 101353158 POLARIS. Opinions, interpretations, conclusions, and recommendations are those of the authors and are not necessarily endorsed by the Department of Defense. We thank Andrew Carlson, David Okonkwo, and Ramon Diaz-Arrastia for helpful discussions. We also thank Jed A. Hartings for providing one of the datasets used in this multicenter study.

